# Prevalence of Elevated Blood Pressure and Left Ventricular Hypertrophy in Adolescents with Congenital Heart Disease

**DOI:** 10.1101/2023.03.15.23287130

**Authors:** Aaron T. Walsh, Kan N. Hor, Mariah Eisner, Mahmoud Kallash, John David Spencer, Andrew H. Tran

## Abstract

**Background:** Left ventricular hypertrophy (LVH) associated with hypertension (HTN) is a predictor of cardiovascular (CV) events in adulthood. LVH is defined using left ventricular mass indexed to height^2.7^ (LVMI-ht^2.7^) with current guidelines using the adult cutoff of 51 g/ht^2.7^; however, the pediatric cutoff is lower. Adults with congenital heart disease (CHD) have higher rates of HTN compared to the general adult population. Data on the prevalence of elevated blood pressure (SBP) in youths with CHD is limited. The aim of our study was to determine the prevalence of elevated BP and LVH in adolescents with CHD.

**Methods:** We retrospectively analyzed echocardiograms from patients with CHD from 2012-2019. Patients with biventricular CHD aged 13-17 years with documented BP, height, weight, and measurement of LVMI-ht^2.7^ were included. We defined LVH using the pediatric cutoff of LVMI-ht^2.7^ ≥ 38.6 g/ht^2.7^. Patients were grouped by BP category into normotensive (NT, SBP < 120 mm Hg), Elevated BP (E-BP, 120 ≤ SBP < 130 mm Hg), Stage 1 HTN (HTN-1, 130 ≤ SBP < 140 mm Hg), and Stage 2 HTN (HTN-2, SBP ≥ 140 mm Hg). Prevalence of LVH was reported in each group defined as LVMI-HT^2.7^ ≥ 38.6 g/ht^2.7^.

**Results:** 855 patients were included. Mean (± standard deviation, SD) age was 15.5±1.5 years with 485/855 (56.7%) male, SBP 117±13.5 mmHg, and LVMI-ht^2.7^ 34.2±10.5 g/ht^2.7^. 493/855 (57.7%) were in the NT group, 214/855 (25%) in E-BP, 99/855 (11.6%) in HTN-1, and 49/855 (5.7%) in HTN-2. Prevalence of LVH increased with higher SBP with 96/493 (19.5%) in NT, 80/214 (37.4%) in E-BP, 32/99 (32.3%) in HTN-1, and 20/49 (40.8%) in HTN-2. Of youths with LVH, 49/228 (21.5%) met adult criteria of ≥ 51 g/ht^2.7^. Age, male sex, and body mass index (BMI) percentile were significantly associated with increased LVMI-ht^2.7^.

**Conclusions:** Youths with CHD have a high prevalence of elevated BP, HTN, and LVH. BMI is a significant risk factor for the development of LVH in this population. These findings support early screening for HTN in this group because youths with CHD have baseline increased CV risk that may be compounded by obesity and long-term HTN.

**Clinical Perspective:**

- What is new?
  - The prevalence of abnormal blood pressure and LVH in adolescents with CHD is understudied and our data show that HTN and LVH are common in this population.
  - One-fifth of adolescents with biventricular congenital heart disease and left ventricular hypertrophy met adult criteria for left ventricular hypertrophy.
- What are the clinical implications?
  - Left ventricular hypertrophy secondary to hypertension is linked to adverse cardiac events in adulthood.
  - Early screening and detection for abnormal blood pressure in the adolescent congenital heart disease population may lead to earlier initiation of lifestyle interventions or pharmacotherapy and mitigate long-term adverse clinical and financial outcomes in an already vulnerable population.

## Introduction

The global prevalence of congenital heart disease (CHD) is estimated to be 1.8 cases per 100 live births^1^ and accounts for one-third of all congenital anomalies^2^. With improvements in prenatal detection, postnatal care, pharmacologic agents, interventional techniques, and surgical outcomes, more patients born with CHD are living past the first decade of life. Adults with CHD now comprise two-thirds of the total CHD population^3^ and manifest several cardiovascular (CV) risk factors at rates higher than the general adult population, including systolic hypertension (HTN)^4^.

Left ventricular hypertrophy (LVH) as detected by echocardiography has long been associated with HTN and confers an increased risk of adverse CV events in adulthood. The Framingham Heart Study demonstrated that increased left ventricular mass (LVM) in adults was independently associated with CV disease, death from CV disease, and all-cause mortality even after adjustment for traditional risk factors^5^. The prognostic ability of LVH for CV events has held true across more diverse ethnic populations, the elderly, in those with essential HTN, and following myocardial infarction^6-12^ with links to increased risk of stroke, heart failure, coronary artery disease, and arrhythmia^6, 7, 12-14^. In the general adolescent population with essential hypertension, a linear relationship was found between HTN and LVM^15^, with LVM values associated with a fourfold increase for CV disease in adulthood^16^. However, the prevalence of HTN and LVH in adolescents with CHD is less well-known. Therefore, the aim of our study was to determine the prevalence of HTN and LVH in this ever-growing population.

## Methods

### Study Population

We performed a retrospective analysis of an echocardiogram database and the Electronic Medical Record of adolescent patients aged 13-17 years with biventricular CHD (excluding those patients with single ventricular or functionally single ventricular lesions such as hypoplastic left heart syndrome, tricuspid atresia, and pulmonary atresia with intact ventricular septum, or others who have required single-ventricle palliation) from 2012-2019 at Nationwide Children’s Hospital. We chose to study this age distribution as the blood pressure cutoffs in this population directly correlate with those in the adult population^17^ in which adverse CV outcomes are described. Only patients with documented blood pressure, height, weight, body mass index (BMI), and echocardiographic data for LVM calculation at the time of their most recent echocardiogram were included.

### Echocardiogram Parameters and LVM

For the purposes of this study, the echocardiographic measurements were measured off M-Mode, a standard for LV quantification in our lab during this timeframe. Demographic data and echocardiographic variables for assessment of LVH are included in **Table 1**. Measurements of the interventricular septum (IVSd) and left ventricular posterior wall (LVPWd) were taken at end-diastole. Left ventricular dimensions were taken at end-diastole (LVEDD) and end-systole (LVESD). LVM was determined via the formula proposed by Devereux et al: 0.8x (1.04x[(IVS+LVID+PWT)^3^-LVID^3^] + 0.6 grams^18^.

**Table 1.** Subject characteristics

| Characteristic | Overall<br>N = 855<sup>1</sup> | Normotensive<br>N = 493<sup>1</sup> | Elevated<br>N = 214<sup>1</sup> | Stage 1<br>HTN<br>N = 99<sup>1</sup> | Stage 2<br>HTN<br>N = 49<sup>1</sup> |
| --- | --- | --- | --- | --- | --- |
| Age (years) | 15.48<br>(1.46) | 15.32 (1.47) | 15.59 (1.51) | 15.91<br>(1.20) | 15.64<br>(1.37) |
| Sex |  |  |  |  |  |
| Female | 364<br>(42.9%) | 243 (49.6%) | 77 (36.3%) | 28 (28.3%) | 16 (33.3%) |
| Male | 485<br>(57.1%) | 247 (50.4%) | 135 (63.7%) | 71 (71.7%) | 32 (66.7%) |
| Race |  |  |  |  |  |
| White | 642<br>(81.9%) | 371 (82.1%) | 156 (79.6%) | 78 (83.9%) | 37 (86.0%) |
| Black | 112<br>(14.3%) | 66 (14.6%) | 30 (15.3%) | 11 (11.8%) | 5 (11.6%) |
| Other | 30 (3.8%) | 15 (3.3%) | 10 (5.1%) | 4 (4.3%) | 1 (2.3%) |
| Height (cm) | 165 (13) | 163 (13) | 168 (13) | 170 (12) | 169 (11) |
| Body surface area (m<sup>2</sup>) | 1.71 (0.31) | 1.62 (0.28) | 1.78 (0.28) | 1.87 (0.28) | 1.96 (0.40) |
| Weight (kg) | 61 (52, 73) | 56 (48, 66) | 66 (56, 78) | 71 (61, 85) | 82 (63, 100) |
| BMI (kg/m<sup>2</sup>) | 22.0 (19.2, 25.9) | 20.7 (18.5, 24.0) | 23.0 (19.9, 26.8) | 24.4 (21.2, 29.3) | 27.3 (21.9, 32.6) |
| BMI percentile (%) | 68 (37, 92) | 60 (29, 84) | 77 (48, 93) | 86 (59, 96) | 95 (78, 98) |
| BMI category |  |  |  |  |  |
| Normal | 570<br>(67.1%) | 377 (76.9%) | 129 (60.8%) | 48 (48.5%) | 16 (33.3%) |
| Overweight | 127<br>(15.0%) | 59 (12.0%) | 37 (17.5%) | 22 (22.2%) | 9 (18.8%) |
| Obese | 152<br>(17.9%) | 54 (11.0%) | 46 (21.7%) | 29 (29.3%) | 23 (47.9%) |
| Systolic blood pressure (mm Hg) | 117 (108, 126) | 110 (104, 114) | 124 (122, 126) | 133 (131, 136) | 146 (142, 152) |
| Diastolic blood pressure (mm Hg) | 64 (59, 70) | 62 (58, 68) | 66 (60, 72) | 67 (60, 76) | 72 (64, 80) |

<sup>1</sup> Mean (SD); n (%); Medians (IQR)
\*HTN, hypertension; BMI, body mass index; SD, standard deviation; IQR, interquartile range

LVM was then indexed to body height to the power of 2.7 (LVMI-ht^2.7^=LVM/height^2.7^) to adjust for differences in body size^18, 19^. LV function was assessed by left ventricular shortening fraction (LVSF).

### Blood Pressure Definitions and Categories

Patients were stratified by blood pressure category into normotensive (NT, SBP<120 mmHg), elevated blood pressure (E-BP, 120≤SBP<130 mmHg), stage 1 HTN (HTN-1, 130≤SBP<140 mmHg), and stage 2 HTN (HTN-2, SBP≥140 mmHg) per the 2017 Clinical Practice Guideline (CPG) for Screening and Management of Blood Pressure in Children and Adolescents^17^ utilizing the automated blood pressure taken at the time of their most recent echocardiogram. The prevalence of LVH was reported in each group using the pediatric cutoff for LVH of LVMI-ht^2.7^ ≥ 38.6 g/ht^2.7^ which corresponds to the 95th percentile for LVMI-ht^2.7^ in children and increases the sensitivity for detecting LVH before it reaches a level associated with CV events in adulthood^15, 20^. Results were also compared to the 2017 CPG recommendation of LVMI-ht^2.7^ ≥ 51g/ht^2.7^ for LVH, which is the adult cutoff associated with increased risk for cardiovascular events ^21^ but which is far above the 95^th^ percentile values in pediatrics ^17^.

### BMI stratification

BMI percentiles were obtained from the Centers for Disease Control and Prevention growth charts. BMI z-scores were calculated by BMIz=(BMI/M)L-1/(L*S), where L, M, and S are reference values from the growth chart for the corresponding child age and sex. BMI percentiles were calculated from z-scores based on the normal distribution. Subjects were categorized based on BMI percentile as normal weight (BMI < 85th percentile), overweight (85th percentile ≤ BMI < 95th percentile), or obese (BMI ≥ 95th percentile).

### Statistical analysis

Data were summarized using frequency (percentage) for categorical variables, mean (standard deviation, SD) for continuous symmetric variables, and median (interquartile range, IQR) for continuous skewed variables. Data were quality checked and distributions were visualized using bar and violin plots. For model stability, race groups were aggregated into white, black, or other (includes American Indian, Asian, Hispanic, Native American, Pacific Islander, biracial, and other). Prevalence of LVH among each blood pressure group was calculated. Among adolescents that met pediatric LVH criteria, the percentage also meeting the adult LVH criteria was calculated. Pairwise comparisons were performed for the echocardiogram parameters between each blood pressure category and adjusted for multiple comparisons utilizing the Bonferroni correction. Univariable linear regression models were constructed for continuous outcomes LVMI-ht^2.7^, LVEDD, LVESD, LVPWd, IVSd, LVSF, and LVEF. Predictor variables included age, sex, race, continuous SBP, and BMI percentile. Multivariable linear regression models were fit for each outcome with the same independent variables, using complete cases without missing predictors. The interaction term between SBP and BMI percentile was also assessed and removed from the model if not significantly associated with the outcome. P-values < 0.05 were considered statistically significant. All statistical analyses were performed in R version 4.0 (R Core Team, Vienna, Austria).

## Results

### Patient Characteristics

A total of 855 patients met the inclusion criteria (56.7% male) with mean (SD) age 15.5 ± 1.5 years (range 13.0 to 17.9 years). Patient characteristics can be found in **Table 1**. The race of patients was predominantly White (79.9%). BMI, BMI percentile, and body surface area (BSA) trended higher in the abnormal blood pressure groups. Over half of patients were NT (57.7%), with 25%, 11.6% and 5.7% meeting criteria for E-BP, HTN-1, and HTN-2, respectively.

### Echocardiogram Parameters

Those in the E-BP and HTN-1 groups had higher LVEDD values compared to NT (**Table 2)**. Patients with abnormal BP had thicker LVPWd and IVSd compared to NT subjects. LVMI-ht^2.7^ increased across blood pressure categories and was significantly higher in the abnormal blood pressure groups compared to NT. One-third of patients in the HTN-1 and 41% of patients in the HTN-2 category met criteria for LVH. Of the youths with LVH, one-fifth (21.5%) met the adult criteria for LVH with LVMI-ht^2.7^ ≥ 51g/ht^2.7^. Ejection fraction was preserved and consistent across blood pressure categories.

**Table 2.**
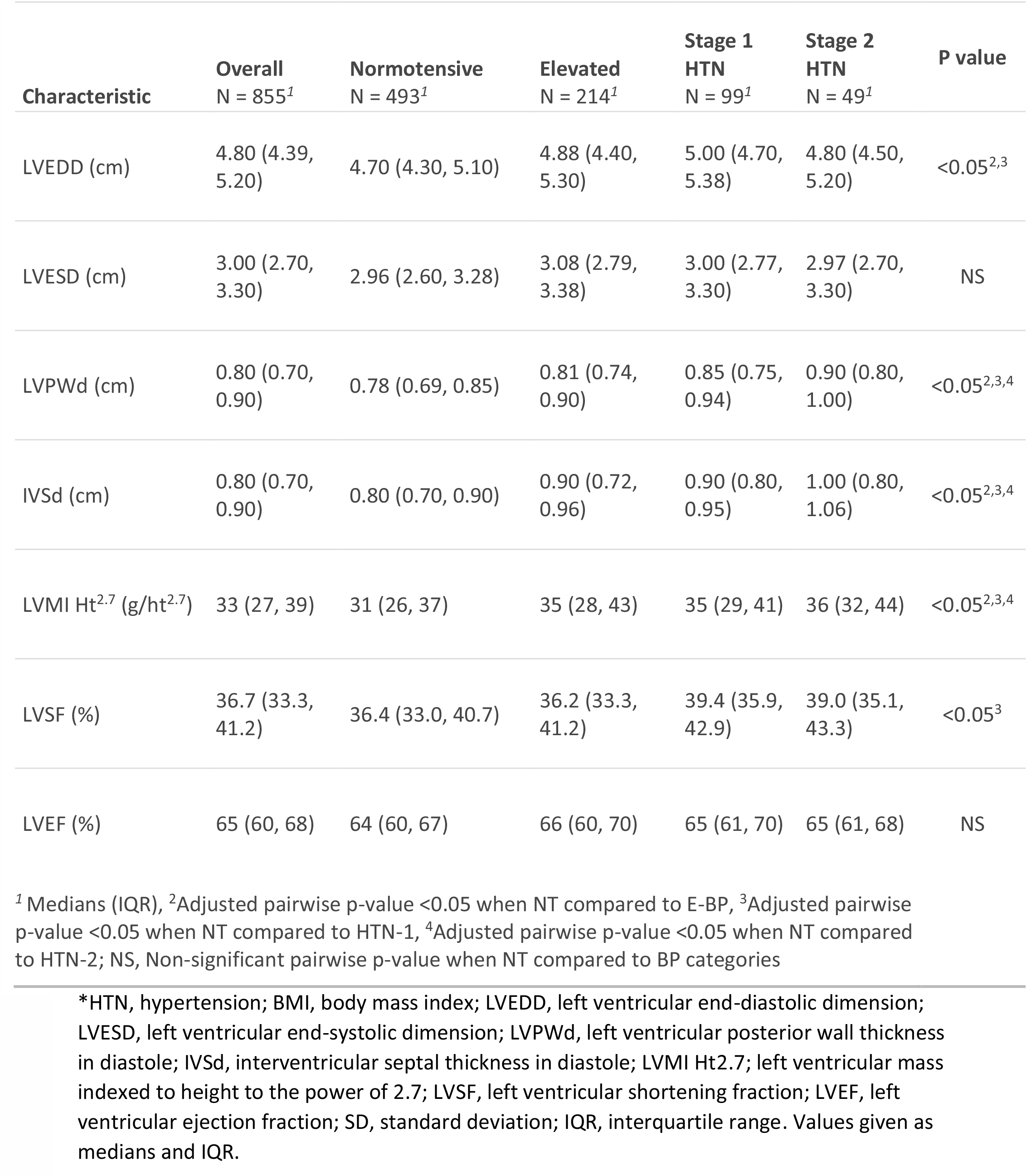
Echocardiogram Parameters

| Characteristic | Overall<br>N = 855<sup>1</sup> | Normotensive<br>N = 493<sup>1</sup> | Elevated<br>N = 214<sup>1</sup> | Stage 1<br>HTN<br>N = 99<sup>1</sup> | Stage 2<br>HTN<br>N = 49<sup>1</sup> | P value |
| --- | --- | --- | --- | --- | --- | --- |
| LVEDD (cm) | 4.80 (4.39, 5.20) | 4.70 (4.30, 5.10) | 4.88 (4.40, 5.30) | 5.00 (4.70, 5.38) | 4.80 (4.50, 5.20) | <0.05<sup>2,3</sup> |
| LVESD (cm) | 3.00 (2.70, 3.30) | 2.96 (2.60, 3.28) | 3.08 (2.79, 3.38) | 3.00 (2.77, 3.30) | 2.97 (2.70, 3.30) | NS |
| LVPWd (cm) | 0.80 (0.70, 0.90) | 0.78 (0.69, 0.85) | 0.81 (0.74, 0.90) | 0.85 (0.75, 0.94) | 0.90 (0.80, 1.00) | <0.05<sup>2,3,4</sup> |
| IVSd (cm) | 0.80 (0.70, 0.90) | 0.80 (0.70, 0.90) | 0.90 (0.72, 0.96) | 0.90 (0.80, 0.95) | 1.00 (0.80, 1.06) | <0.05<sup>2,3,4</sup> |
| LVMI Ht<sup>2.7</sup> (g/ht<sup>2.7</sup>) | 33 (27, 39) | 31 (26, 37) | 35 (28, 43) | 35 (29, 41) | 36 (32, 44) | <0.05<sup>2,3,4</sup> |
| LVSF (%) | 36.7 (33.3, 41.2) | 36.4 (33.0, 40.7) | 36.2 (33.3, 41.2) | 39.4 (35.9, 42.9) | 39.0 (35.1, 43.3) | <0.05<sup>3</sup> |
| LVEF (%) | 65 (60, 68) | 64 (60, 67) | 66 (60, 70) | 65 (61, 70) | 65 (61, 68) | NS |

<sup>1</sup> Medians (IQR), <sup>2</sup>Adjusted pairwise p-value <0.05 when NT compared to E-BP, <sup>3</sup>Adjusted pairwise p-value <0.05 when NT compared to HTN-1, <sup>4</sup>Adjusted pairwise p-value <0.05 when NT compared to HTN-2; NS, Non-significant pairwise p-value when NT compared to BP categories
\*HTN, hypertension; BMI, body mass index; LVEDD, left ventricular end-diastolic dimension; LVESD, left ventricular end-systolic dimension; LVPWd, left ventricular posterior wall thickness in diastole; IVSd, interventricular septal thickness in diastole; LVMI Ht<sup>2.7</sup>; left ventricular mass indexed to height to the power of 2.7; LVSF, left ventricular shortening fraction; LVEF, left ventricular ejection fraction; SD, standard deviation; IQR, interquartile range. Values given as medians and IQR.

### Univariable and Multivariable Linear Regression Models

Univariable linear regression demonstrated age, male sex, BMI percentile, and SBP were significantly associated with LVH **(Table 3)**. After adjustment in a multivariable linear regression, age, male sex, and BMI percentile remained significant predictors of LVMI-ht^2.7^ while SBP no longer correlated **(Table 4)**. In a subgroup analysis of patients with normal BMI, SBP was an independent predictor of LVMI-ht^2.7^ (p=0.011) on univariable analysis, but lost significance in the adjusted model.

**Table 3.**
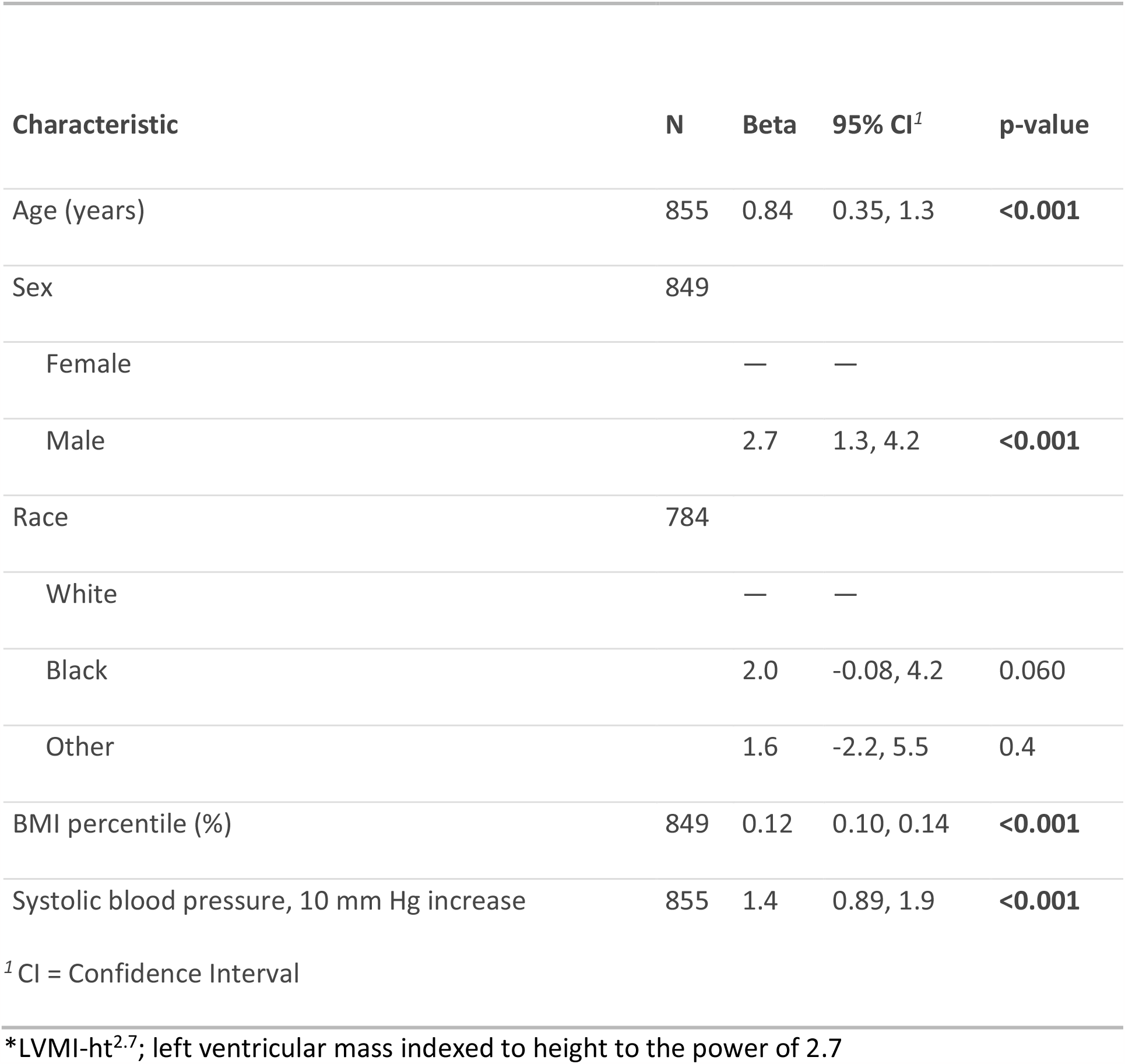
Univariable linear regression model for LVMI-ht^2.7^

| Characteristic | N | Beta | 95% CI<sup>1</sup> | p-value |
| --- | --- | --- | --- | --- |
| Age (years) | 855 | 0.84 | 0.35, 1.3 | **<0.001** |
| Sex | 849 |  |  |  |
| Female |  | — | — |  |
| Male |  | 2.7 | 1.3, 4.2 | **<0.001** |
| Race | 784 |  |  |  |
| White |  | — | — |  |
| Black |  | 2.0 | -0.08, 4.2 | 0.060 |
| Other |  | 1.6 | -2.2, 5.5 | 0.4 |
| BMI percentile (%) | 849 | 0.12 | 0.10, 0.14 | **<0.001** |
| Systolic blood pressure, 10 mm Hg increase | 855 | 1.4 | 0.89, 1.9 | **<0.001** |
| <sup>1</sup> CI = Confidence Interval |  |  |  |  |

\*LVMI-ht<sup>2.7</sup>; left ventricular mass indexed to height to the power of 2.7

**Table 4.** Multivariable linear regression model for LVMI-ht^2.7^

| Characteristic | Beta | 95% CI<sup>1</sup> | p-value |
| --- | --- | --- | --- |
| Age (years) | 0.77 | 0.30, 1.2 | **0.001** |
| Sex |  |  |  |
| Female | — | — |  |
| Male | 2.7 | 1.3, 4.1 | **<0.001** |
| Race |  |  |  |
| White | — | — |  |
| Black | 1.7 | -0.21, 3.7 | 0.080 |
| Other | 1.9 | -1.7, 5.5 | 0.3 |
| BMI percentile (%) | 0.11 | 0.09, 0.14 | **<0.001** |
| Systolic blood pressure, 10 mm Hg increase | 0.26 | -0.29, 0.81 | 0.4 |

\*LVMI-ht<sup>2.7</sup>, left ventricular mass indexed to height to the power of 2.7; BMI, body mass index

## Discussion

### HTN

There is a paucity of data related to HTN and LVH in the adolescent congenital heart disease population. We found that in our large cohort of adolescents with biventricular CHD, over 40% had abnormal blood pressure. LVM was linearly associated with elevated blood pressure such that only 20% of those with normal blood pressure had LVH by echocardiography compared to 35% of those with HTN. The rates of abnormal blood pressure in our group were higher than those previously reported for youth in the general population^22-26^, which may reflect the impact of the underlying CHD on vascular tone. Indeed, Mivelaz et al demonstrated elevated aortic stiffness in a similar cohort of adolescents with biventricular CHD^27^, which is a vascular marker linked to future cardiovascular morbidity and mortality^28^. This is not surprising given that patients with CHD often have several risk factors for HTN including chronic renal insufficiency as well as changes in arterial wall architecture that places them at higher risk of abnormal blood pressure^29, 30^. High blood pressure did not remain a significant risk factor for LVH in multivariable regression analysis. We suspect this may be due to a longitudinal effect of blood pressure on LVM that may not be seen until the adult years. Nonetheless, we know that hypertensive youths are prone to becoming adults with HTN ^31-33^ and that identification of abnormal blood pressures in the formative years may play an important prognostic role.

Perhaps annual ambulatory blood pressure monitoring in adolescents with CHD would be best practice to screen for those already experiencing elevated blood pressures.

### LVH

LVMI-ht^2.7^ appeared to be higher in those with higher BP on univariate analysis consistent with previous findings in children and adolescents without CHD^34^. One-fifth of patients with LVH met adult criteria which is higher than levels previously reported for hypertensive young patients without CHD^16^, suggesting a compounding effect of underlying CHD on the development of LVH. Age, male sex, BMI percentile, and blood pressure were significantly associated with LVMI-ht^2.7^ using univariable analysis. After adjusting for variables in a multilinear regression model, age, male sex, and BMI percentile remained significant predictors of LVMI-ht^2.7^ while SBP lost its significance, similar to previous findings in adults by de Simone et al. ^18^. However, data from the SHIP AHOY study group have demonstrated SBP to be a significant determinant of LVMI-ht^2.7^ in healthy adolescents after adjusting for covariates^15^. Their cohort age range skewed older than the current data set, suggesting a longitudinal effect of SBP on LVMI-ht^2.7^ which may have risen to significance on multivariate analysis in our cohort if given more time.

### BMI

The association between obesity, HTN, and LVH in the general pediatric population is well-documented^35-38^. Children without CHD with overweight or obesity and HTN were found to have higher LVM indices at baseline and longitudinally compared to those of healthy weight who were hypertensive^39^. Our study of children with CHD mirrors these findings with one in five patients having obesity and BMI percentile being significantly associated with LVMI-ht^2.7^. This is important because children with CHD can be at higher risk for future CV events compared to non-CHD children^40^ so the presence of obesity and early echocardiographic changes may further compound future risk in these patients.

Weight continues to prove itself to be a crucial factor in the development of LVH. Similar to our findings, in a cohort of 160 pediatric patients from the Bogalusa Heart study, weight remained the only significant predictor for LVM in multivariate analysis after controlling for age, sex, and SBP ^41^. Burke et al. found that adults in the MESA study who were obese had a two-fold increased risk of having LVM exceeding the 80^th^ percentile compared to those with a normal body size after adjustment for traditional risk factors, making body size a potentially modifiable risk factor for subclinical CV disease^42^.

Our study was novel in having a large cohort of patients inclusive of a large spectrum of biventricular CHD. We also utilized the most up to date guidelines for characterizing HTN in the adolescent population^17^ which is felt to capture more accurately those with abnormal blood pressures. Prior studies on HTN and LV geometry changes in CHD have focused primarily on coarctation of the aorta and its associated long-term outcomes^43, 44^. While our study included patients with left ventricular outflow tract (LVOT) obstructions, the majority of documented LVOT gradients on the echocardiograms used for this study demonstrated no or mild residual pressure gradient, indicating that a residual obstruction is less likely responsible for any observed increase in LVM. This study broadens our understanding of the impact of HTN and overweight/obesity on children with CHD. Our study highlights the need for early detection of abnormal BP and obesity in patients with CHD to prevent progression of LVH to levels known to be associated with hard CV events. Knowledge of the risk for LVH in this age group becomes increasingly important considering findings that elevated LVM can be found at blood pressures below the current definition of pediatric HTN ^15^. Furthermore, elevated blood pressure has been associated with systolic and diastolic dysfunction^45, 46^ which may already be compromised by the patient’s CHD. With the knowledge of early abnormal blood pressure and evidence of increased LVM in overweight and obese patients with CHD, it becomes vital to screen patients and to be vigilant about providing anticipatory guidance and considering pharmacological intervention in patients who meet criteria. This is especially important in light of evidence that regression in LVM leads to lesser risk of CV disease ^8^. Targeted diet and lifestyle recommendations such as the Dietary Approaches to Stop Hypertension (DASH) ^17^ and the Cardiovascular Health Integrated Lifestyle Diet (CHILD) recommended by the American Academy of Pediatrics (AAP) ^47^ should be stressed particularly in this patient population.

Physical activity recommendations should also be emphasized ^48^ depending on appropriate activity clearance by heart lesion. Current pediatric hypertension guidelines recommend yearly ambulatory blood pressure monitor (ABPM) placement in patients with repaired aortic coarctation ^17^; however, ABPM can also be useful in screening other CHD patients for hypertension.

### Limitations

While our study is one of the largest to evaluate BP and LVMI-ht^2.7^ amongst the pediatric CHD population, there are several inherent limitations that should be noted. Our study is a single center retrospective cohort study which limits the ability to determine causation for increased LVM. We believe that with our large sample size, which encompasses all types of biventricular CHD, we avoided selection bias that can accompany some retrospective studies. To that end, we did not stratify by types of CHD and it is possible that our population potentially includes a larger percentage of patients with left ventricular outflow tract obstructive lesions which are known to be risk factors for LVH. This will need to be further elucidated in future studies. Given the descriptive nature of our work, whether patients were identified as hypertensive or if they were being actively treated for HTN was not investigated. Additionally, single time-point blood pressures were utilized for our analysis. Serial blood pressure evaluation and incorporation of ambulatory blood pressure monitoring data would further enhance our understanding of the true prevalence of HTN in this cohort and provide insight into the timing of development of end-organ damage. Future studies should be aimed at longitudinal changes in blood pressure and LVH and the impact of pharmacological intervention and social determinants on HTN, obesity, and evidence of cardiac dysfunction in the pediatric CHD population.

## Data Availability

All data produced in the present study are available upon request to the authors.

## Acknowledgements

Sources of Funding: None

